# Assessing EHR potential for adaptive learning in multimorbidity care in Sub-Saharan Africa: a mixed-methods study of Zimbabw’s Impilo system

**DOI:** 10.64898/2026.07.16.26357920

**Authors:** Efison Dhodho, Kety Choga, Fiona Mundoga, Pugie T Chimberengwa, Robert Tawanda Gongora, Trymore Chaurura, Karen Webb, Memory Benjamin, Theonevus T Chinyanga, Forget Banda, Kenneth Masiye, Nicholas Midzi, Justice Mudavanhu, Agnes Katsidzira, Trudy Mhlanga, Patience Mangisi, Tsitsi Apollo, Clorata Gwanzura, Valiant Makore, Shingirirayi Tsvangirayi, Cleopas Chimbetete, Blessing Manyiyo, Justin Dixon, Dorothea Nitsch

## Abstract

Electronic health records (EHR) are increasingly recognised as critical digital infrastructure for integrated, patient-centred care in the context of rising multimorbidity. In low-resource settings, national EHRs may also support locally driven learning to improve adaptive care across chronic conditions. However, there is limited empirical evidence on whether and how these systems enable learning within routine care in ways that inform broader system adaptation. We conducted a qualitative multi-method assessment of Impilo, Zimbabw’s national EHR, to examine its capacity to support learning for integrated multimorbidity care at primary care level, using HIV-hypertension as a tracer condition pair. Guided by Friedman’s socio-technical infrastructure model as the analytical framework and Learning Health Systems (LHS) theory as the interpretive framework, data were drawn from documentary review, ethnographic observation, patient journey mapping, and interviews with frontline health workers and key stakeholders.

Frontline learning for person-centred multimorbidity care was actively generated through interpretation of patient trajectories, experiential adjustment, and coordination across HIV and hypertension services using both the EHR and paper-based artefacts such as registers and patient booklets. However, this learning remained largely encounter-bound and weakly stabilised. Impilo did not routinely provide usable longitudinal patient views, practice-facing analytic tools, or institutionalised mechanisms for collective reflection required to support integrated multimorbidity care. Consequently, learning was largely confined to incremental adjustment within existing workflows, with limited capacity to inform broader changes to care pathways, routines, or system design.

These findings suggest that the principal barrier to developing LHS is not the absence of data or frontline learning capacity, but the lack of socio-technical arrangements that enable learning to stabilise and inform system adaptation. Digitalisation alone is insufficient to support adaptive multimorbidity care. Co-production with frontline health workers may provide a pathway for aligning digital system design with routine care realities.

**Author summary:** Electronic health records are increasingly used to support integrated care for people living with multiple long-term conditions, including HIV and hypertension. However, they practically remain organised around single diseases and reporting requirements, limiting their ability to support coordinated, person-centred care. We studied Zimbabw’s national electronic health record system, Impilo, to understand how it supports integrated HIV and hypertension care in primary healthcare settings. We found that frontline health workers routinely generated practical knowledge by interpreting patient histories, coordinating care across services, and combining paper and electronic records to maintain continuity of care. However, much of this learning remained informal and tied to individual patient encounters because the system did not routinely provide longitudinal patient views, coordinated follow-up, or integrated information across conditions. Our findings suggest that the main challenge is not a lack of data, digital systems, or frontline expertise, but the limited ability of current systems to support learning and adaptation for multimorbidity care. Involving frontline health workers in the design and improvement of electronic health records may help create more integrated, learning-oriented health systems.

## 1. Introduction and Background

In low-resource settings experiencing rapid epidemiological transition, non-communicable diseases (NCDs) are increasingly prevalent(1) and often co-occur with persisting chronic infectious conditions such as HIV (2). Across Africa, this convergence has led to rising multimorbidity—the coexistence of ≥2 chronic conditions—which has emerged a critical pressure point for many health systems in the region(3). Zimbabwe reflects these trends. Among its large, ageing population of people living with HIV (PLHIV), HIV-hypertension has become the most prevalent and pressing multimorbidity cluster(4). Recent large-scale screening of over 120,000 individuals across four districts revealed that 48% of PLHIV also had hypertension. (5)

Growing recognition of multimorbidity has accelerated initiatives to deliver integrated, person-centred chronic care, widely thought to be necessary for responding to this complexity (6). Within such pushes, digital health interventions - including electronic health records (EHRs), telehealth, mobile applications, and remote monitoring (7) - are expected to support decentralisation and task-sharing, integrated decision-support, longitudinal follow-up, and improved service delivery. However, despite growing investment in digital health infrastructure across Africa, health systems remain unable to effectively utilize these tools to meet the complex needs of patients living with chronic conditions and multimorbidity (8,9). More data are being collected, yet little of this information is meaningfully used to support care and improvement at the point of service delivery(10,11).

Our previous work in Zimbabwe has reframed multimorbidity not simply as a clustering of diseases but as a manifestation of a deeper structural mismatch between rigid, disease-centred policies and guidelines within vertically organised health systems and the complex, evolving realities of patient care (12). From this perspective, multimorbidity exposes a more fundamental challenge: health systems struggle to learn from and adapt to changing patterns of disease and care needs (12). This challenge is most acute at the frontline, where health workers and patients themselves must respond to increasing complexity while managing long-term care with limited resources(13). Despite incremental, programme-driven efforts integrate HIV and NCD services over the last decade, patients continue to navigate fragmented pathways, often within the same facilities(14). Digital tools that could support integration and ease this burden remain underutilised, because they are not designed to support reflection, coordination, and decision-making in everyday clinical practice (2,15). These challenges are further compounded by externally funded, vertically-structured disease control programmes, which prioritise reporting and upward accountability over local learning and adaptation(2,12).

LHS offer a conceptual horizon for health system transformation. Although the concept originated in high-income settings, it is increasingly relevant for African health systems confronting complexity, fragmentation, and extractive knowledge systems that often undermine meaningful and sustainable integration.(11,16). Elements of these principles have already been practised, often through quality improvement initiatives implemented at facility, district, or programme level, though not always recognised or developed as formal LHS approaches. LHS are characterised by the continuous generation and use of knowledge to improve services and system functions through embedded feedback loops, collective reflection, and iterative adaptation (11,17). They depend on accessible, real-time data and, by extension, on information systems such as EHR (18). While national digital health strategies increasingly position EHRs as foundational to LHS, there remains limited empirical evidence on whether—and how— existing national EHRs in low- and middle-income countries support cumulative, deliberative learning in routine care(19,20).

For EHRs to support learning, they must do more than store clinical data. Foundational LHS scholarship conceptualises digital infrastructure as a core enabler of continuous improvement rather than merely a repository of patient records (21). Similarly, one of the most significant systematic review by of LHS Enticott et al, upon which this research draws, reached the conclusion that EHR-enabled learning depends on the ability to transform routine clinical data into actionable knowledge that can inform care and system improvement (16). To support such learning, EHRs must enable whole-person patient views, longitudinal patient trajectories, cross-condition visibility, coordinated care and follow-up, and information that supports reflection and continuous improvement. The importance of whole-person representations is well established within patient-centred care and EHR design literature, which argues that digital systems should support understanding of patients as whole persons rather than collections of discrete conditions(22). Likewise, integrated care frameworks emphasise the need for longitudinal information sharing, care coordination, and visibility across providers and conditions to support continuity of care (23). However, while these functions are discussed across the LHS, patient-centred care, integrated care, and digital health literatures, they are rarely considered collectively as the informational foundation required for learning-oriented multimorbidity care. Consequently, there remains limited empirical understanding of whether existing national EHR systems in low-resource settings possess these capabilities, and how their presence or absence shapes frontline learning and service delivery.

In Zimbabwe, national investment in digital health has increased in recent years, most notably through the development of the EHR (15,24). Using HIV-hypertension multimorbidity as a critical test case, this study assesses whether and how Impilo enables or constraints the emergence of sustained structured learning processes in everyday clinical practice. Specifically, it asks: Is learning to improve multimorbidity care possible within this system, and why? Rather than treating Impilo as a standalone software artefact, this study situates it within the broader socio-technical system, linking infrastructural capacity to frontline learning dynamics. In doing so, it conceptualises EHRs not simply as reporting tools, but as core infrastructure for learning. While grounded in Zimbabwe, the challenges identified reflect wider patterns across African health systems seeking to move from data accumulation to adaptive, learning-oriented care (14).

This study addresses these questions through three main objectives:

1. To analyse national health information strategies, policies, and governance arrangement, and assess how the conceptualisation, design, and deployment of the Impilo EHR enable or constrain integrated multimorbidity care.
2. To examine the technical architecture, operational ecosystem, and everyday use of the Impilo EHR at the frontline focusing on data capture, usability, analytic support, and feedback mechanisms for multimorbidity management.
3. To evaluate the extent to which the Impilo EHR enables or inhibits the core functions of a LHS at the point of care in the context of person-centred HIV-hypertension multimorbidity.

## 2. Materials and Methods

### 2.1 Study Design

This mixed-methods study addressed each of the above objectives using:

1. A targeted review of context-specific empirical research, national policies and technical documents shaping health information systems, including Impilo (Objective 1);
2. A structured appraisal of Impilòs technical architecture and everyday use at the point of care using ethnographic patient journey mapping and wider facility observation (Objectives 2 and 3); and
3. Semi-structured interviews with system actors spanning service delivery, management, and technical oversight (Objectives 2 and 3).

Together, these components enabled triangulation between formal system design, technical functionality, and lived experience of system use. This study is reported in accordance with the Standards for Reporting Qualitative Research (SRQR) guideline. Given the multi-method qualitative design incorporating documentary review, ethnographic observation, patient journey mapping, and semi-structured interviews, SRQR was considered the most appropriate reporting framework (25). The completed SRQR checklist is provided as Supporting Information (S1 Checklist).

Reflexivity was considered throughout the study. Several members of the research team had prior professional relationships with participants through their involvement in national digital health, health systems strengthening, and multimorbidity initiatives in Zimbabwe. The multidisciplinary team included clinicians, health systems researchers, epidemiologists, digital health practitioners, and implementation scientists. Regular analytic discussions were used to reflect on how these professional backgrounds and existing relationships might influence data collection, interpretation, and conclusions.

### 2.2 Study Setting

Data collection aimed to take a whole-health system perspective to capture the multidimensional role of Impilo EHR but was empirically anchored in two urban public-sector “learning facilities”: one in Chitungwiza and one in Bulawayo. Like all primary care services in Zimbabwe, both are nurse-led, doctor-supported primary clinics. Fig 1

**Figure 1:**
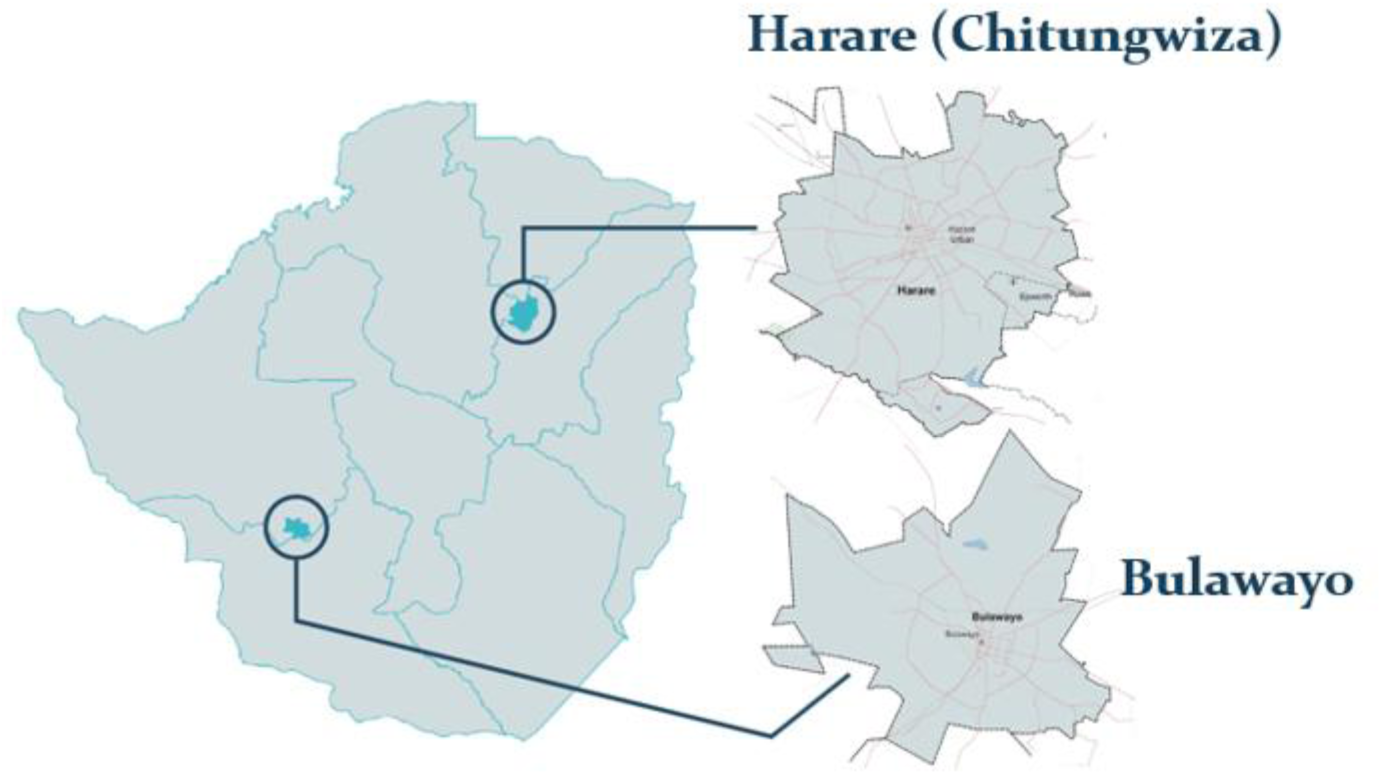
OptiMuL study settings and primary care learning sites in the metropolitan provinces of Harare and Bulawayo.

While recent national guidance has been developed to promote integration of NCD services for PLHIV, both clinics retain the legacy of standalone HIV treatment units (known as ‘opportunistic infection’(OI) departments) and outpatient departments (OPD) where NCDs are seen. Clinic 1 (Chitungwiza) had, in addition to an OPD, a dedicated ‘chronic clinic’ where patients established on treatment for NCDs were seen; in Clinic 2 (Bulawayo), patients with NCDs were seen in the general OPD. These facilities selected on the basis of being among the earliest adopters of the national EHR (26), and are classified as ‘EHR-optimized’ sites, as well as being ones that had taken part in recent NCD screening initiatives (5).

#### Impilo EHR

Impilo is the MoHCC flagship, standards-based, interoperable national EHR, conceived to replace fragmented paper-based registration, clinical documentation and reporting with a unified patient-centric record that supports continuity of care, routine programme monitoring, and surveillance initially for HIV and TB epidemic control, while strengthening the timeliness and quality of strategic information for decision-making. In theory, it is ideally placed to support integrate, adaptive learning to improve multimorbidity care. Its development began in 2015, with an initial pilot in 2016 (emerging in part to address the limitations of the earlier HIV-focused EPMS, including lack of inter-facility connectivity and the inability to assemble a deduplicated whole-patient record), and implementation for national scale-up commenced on 29 September 2022 (15). National scale-up has been supported through PEPFAR financing alongside explicit Government of Zimbabwe co-investment and stewardship, signalling substantial domestic ownership of the platform. The platform has expanded modularly, beginning with “core” workflow components (e.g., client registration and general consultation) and priority HIV/TB functionality (HIV testing services and HIV care and treatment/ART, plus TB), then broadening into wider PHC and MCH service areas; by version 1.3 it comprises 27 modules, with 23 in production (including the Chronic Care module) and additional modules sequenced for deployment across 2025-2026 (19). As of 2025, Impilo had been deployed to 1,254 facilities across all ten provinces, reflecting both substantial reach and the ongoing, phased nature of full digitisation. The current chronic module which is under the Outpatients (Fig 2), does not yet have sophisticated longitudinal patient management features especially when compared to the ART modules and it also does not cater for multiple chronic conditions.

**Fig 2.**
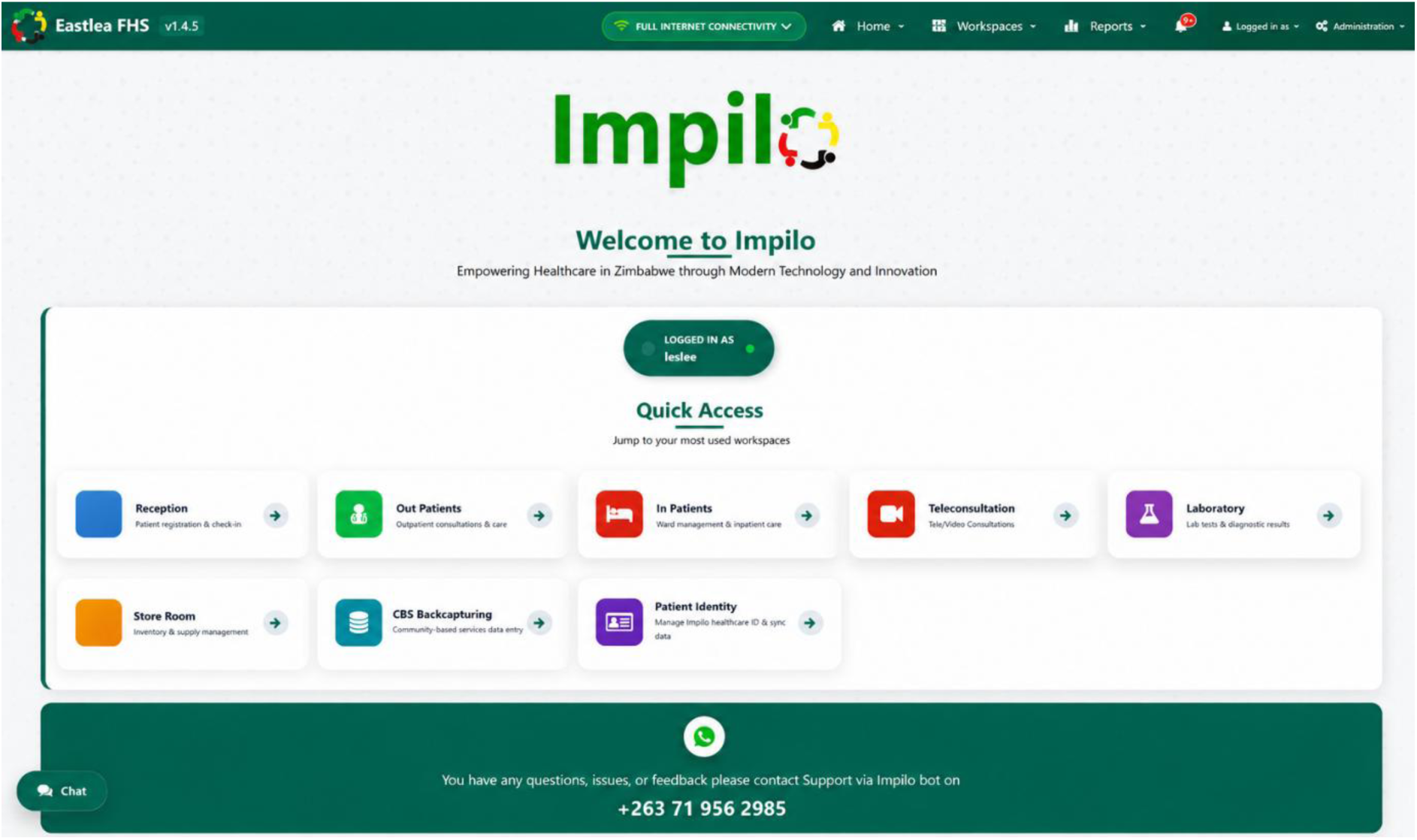
Screenshot of the Impilo electronic health record landing page illustrating the patient management interface used within Zimbabw’s national health information system.

Compared with many LMIC digital health programmes that remain fragmented across disease-specific platforms, pilot implementations, or vendor-dependent architectures, Impilo represents a comparatively ambitious attempt to establish a nationally coordinated, standards-based and government-owned EHR infrastructure at scale (15,20). In this respect, Zimbabwe has charted a relatively distinctive pathway in Sub-Saharan Africa through sustained Ministry stewardship, national interoperability ambitions, and progressive modular expansion within the public health system(19). However, as observed in several donor-supported digital health transitions in LMICs, implementation priorities have remained strongly shaped by vertical HIV programme imperatives and reporting needs (2). While this orientation accelerated national scale-up, it also meant that longitudinal multimorbidity management and cross-condition learning functionalities evolved more slowly than HIV programme capabilities.This combination of strong national infrastructure ambition alongside persistent programme-specific optimisation makes Impilo an important case through which to examine the conditions under which national EHRs can, or fail to, function as infrastructures for LHS in multimorbidity care.

### 2.3 Participant selection

Participants were purposively selected to capture diverse, information-rich perspectives across the socio-technical ecosystem of digital health and multimorbidity care, including actors involved in policy, implementation, clinical delivery, and system governance (28) Consistent with Palinkas et al., this approach enabled the deliberate inclusion of participants with specific knowledge, experience, and roles considered critical for understanding the co-production and implementation of an EHR-supported LHS within complex real-world settings. Building on a prior situational analysis of multimorbidity preparedness in Zimbabwe (29),participants were included across four groups: facility-based actors (nurses, clinicians, data clerks, and facility managers); district- and national-level programme personnel responsible for HIV, NCDs, and digital health governance; technical partners involved in the design and deployment of Impilo; and expert patients living with multimorbidity who interacted with Impilo-supported services. Inclusion was restricted to individuals directly involved in EHR use, governance, or multimorbidity care, including expert patients. Individuals outside these domains and those under 18 years of age were excluded.

### 2.4 Conceptual framework

This study was guided by a two-panel conceptual framework (Fig 2) that enabled assessment of whether Zimbabwe Impilo EHR, considered within its broader NHIS ecosystem, possesses the socio-technical capabilities required to support learning-oriented multimorbidity care. The framework was developed in response to growing recognition that effective multimorbidity care depends on the ability of health information systems to capture, generate, integrate, and process patient information to support continuous adaptation of clinical practice and service delivery.

**Panel A** presents the assessment framework developed for this study. We adapted Friedman et al.’s socio-technical infrastructure model for LHS (18). Within this perspective, we conceptualised Impilo EHR not simply as a repository of patient data, but as part of a socio-technical infrastructure capable of enabling learning through the generation and use of routine health information. We selected Friedman’s framework because the study sought to assess whether the foundational capabilities required for learning exist within the health information system, rather than evaluate implementation success, technology adoption, or routine information system performance. While alternative frameworks such as HOT-Fit and PRISM have been widely applied in digital health and health information systems asessment, they focus primarily on organisational fit, system utilisation, and determinants of data use (30,31). In contrast, we saw that Friedman’s framework explicitly conceptualises the socio-technical services required to support learning, making it particularly suited to our objective of examining the infrastructural foundations of learning-oriented multimorbidity care.

We selected five of Friedman’s ten infrastructural domains aligned to digital infrastructure required for learning to occur: (i) capturing practice as structured data, (ii) storing information in usable repositories, (iii) governing and enabling access to information, (iv) transforming data into actionable knowledge, and (v) returning knowledge to practice. These five domains formed the primary analytical lens through which policy documents, strategic plans, technical artefacts, and frontline clinical practices were examined. The assessment focused on whether Zimbabw’s Impilo EHR possesses the capabilities required to generate information that supports multimorbidity care, including whole-person patient views, longitudinal patient trajectories, cross-condition visibility, coordinated care and follow-up, and data-driven clinical decision-making.

**Panel B** presents the interpretation framework and is adopted from Dixon et al.(2) under CC BY licence, which builds on the LHS framework developed by Sheikh and Abimbola through the World Health Organisation Alliance for Health Policy and Systems Research (17). We selected this framework because we sought not only to assess whether infrastructure capable of supporting learning exists, but also to understand how learning emerges within routine multimorbidity care. Unlike Friedman’s framework, which focuses on the socio-technical prerequisites for learning, the WHO LHS framework provides an interpretive lens for examining how information is translated into deliberation, action, and adaptation within health systems. Drawing on LHS theory, the framework conceptualises learning as the interaction between information, deliberation, and action across single-, double-, and triple-loop learning processes. It therefore provided us with the interpretive scaffolding to assess whether information generated through Zimbabw’s health information infrastructure could support learning and adaptation at clinical, organisational, and system levels

Taken together, Panels A and B provide a structured approach for examining the relationship between digital health infrastructure and learning-oriented multimorbidity care. Building on evidence that a small set of information artefacts can have disproportionate value in translating data into action at the frontline(21,32,33), we incorporated these outputs as a conceptual bridge linking the two panels. The bridge represents what we termed learning-oriented information capability—the informational conditions through which socio-technical infrastructure may enable health system learning. In this study, the concept is operationalised through five outputs: whole-person patient views, longitudinal patient trajectories, cross-condition visibility, coordinated care and follow-up, and data-driven decision-making for continuous improvement (16). It therefore provides the conceptual link between the socio-technical capabilities assessed in Panel A and the learning processes interpreted through the LHS framework in Panel B.

### 2.5 Data sources and data collection

Four primary sources of data (documentary review, Semi-structured interviews, ethnographic patient journey maps and participant observation) are used to assess the readiness of Zimbabw’s Impilo within its broader digital health infrastructure. Data were collected by a team that included a clinical academic and strategic information specialist (ED), a medical anthropologist (JD), a junior social scientist (FM), and research assistant (KC), supported by all other authors. Table 1 below summarizes the data collection and characteristics of respondents.

**Table 1:**
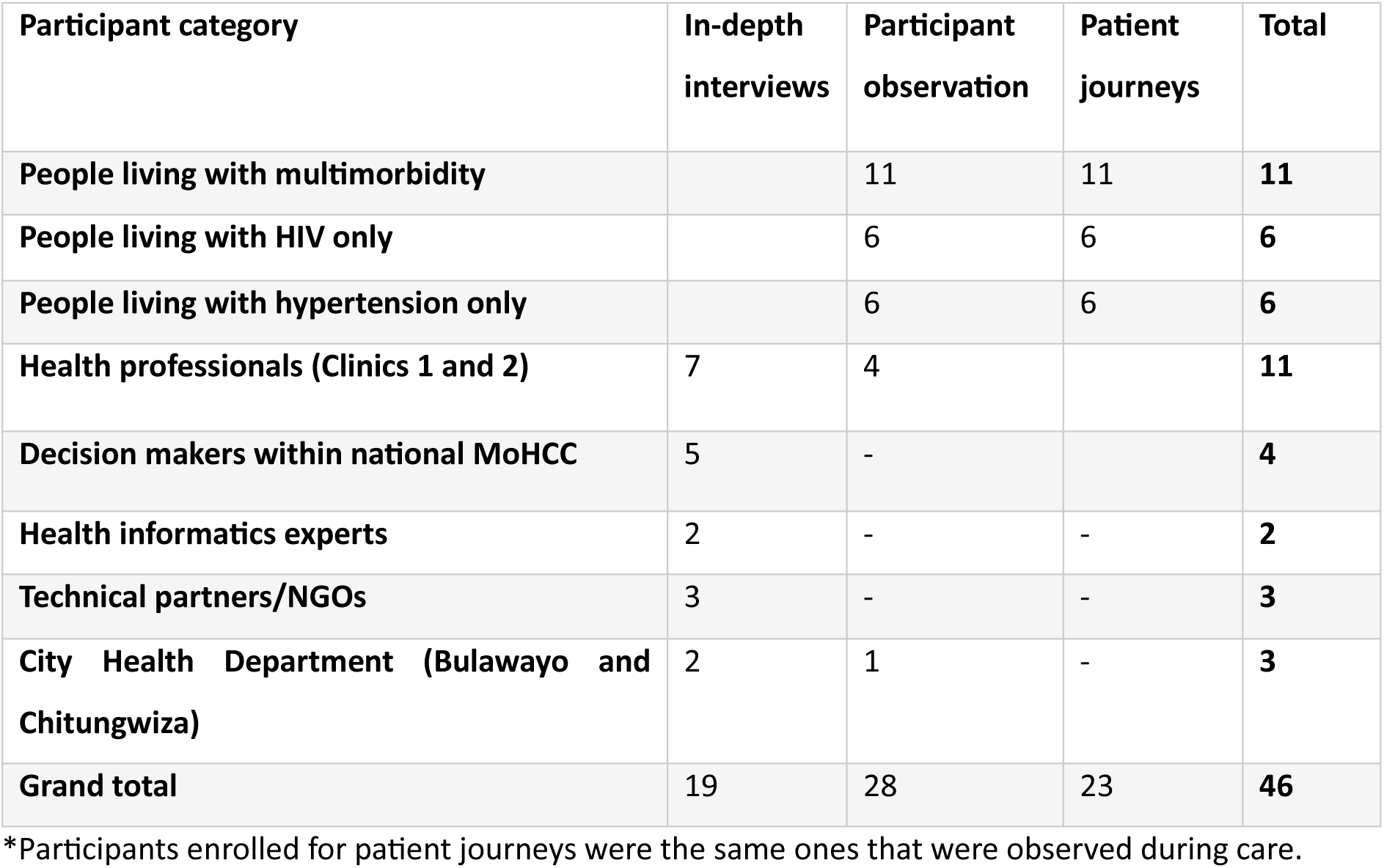
Participant demographics and activity breakdown.

#### 2.4.1 Document and policy review

We conducted a documentary review of national digital health policies, strategies, and technical documents relevant to the design and implementation of the Impilo EHR and the management of multimorbidity in Zimbabwe. The review followed the READ (Ready materials, Extract data, Analyse data, Distil) approach for desk-based document analysis (34). The READ approach was appropriate because it provides a structured yet flexible method for systematically interrogating policy and technical documents to generate contextual insights relevant to complex health systems and implementation processes and at the same time enabled the critical trustworthiness criteria(34).

We identified documents between March 2018 and December 2025 from the Ministry of Health and Child Care, technical and implementing partners, and academic sources known to have influenced multimorbidity/EHR policy and thinking in Zimbabwe. An initial pool of over 40 documents was screened for relevance, resulting in a final analytic set of 13 documents selected for in-depth analysis. These documents were categorised into digital health core documents, programme and clinical guidance, system-level governance materials, and empirical research (S2 Table). Selected EHR validation workshop reports were included to examine how register-validation processes informed EHR design and release decisions. To support transparency and rigour, documents were managed using an agreed file-naming convention, and decisions regarding document inclusion and categorisation were discussed through peer debriefing and member checking with senior researchers, consistent with recommended approaches to enhancing trustworthiness in qualitative document analysis(35).

#### 2.4.2 System appraisal through patient journey and facility ethnography

We conducted a bottom-up system appraisal of the Impilo EHR to examine how its architecture and functionality supported the management of patients with multimorbidity. Following written informed consent, we undertook patient journey mapping (n=23), and concurrent facility-based observation at Clinic 1 and Clinic 2. Patient journeys were mapped by shadowing consenting patients (and, in one case, a treatment supporter) as they navigated routine service delivery points, including registration, clinical consultation, medication dispensing, and follow-up scheduling. Patient journey mapping was selected because it enables detailed examination of how patients, providers, and information systems interact across care pathways, making visible temporal delays, workflow fragmentation, and gaps in continuity of care that are often obscured in conventional workflow assessments(36). We utilized the service blueprinting approach(37) to construct time-stamped, three-lane maps documenting the patient experience, frontline clinical workflow, and associated data flows simultaneously, enabling examination of how clinical encounters and EHR documentation intersected in routine multimorbidity care(see fig 3). Observation durations ranged from approximately two to seven hours per journey, allowing documentation of both single-service visits and more complex, multi-service care trajectories.

**Fig 3.**
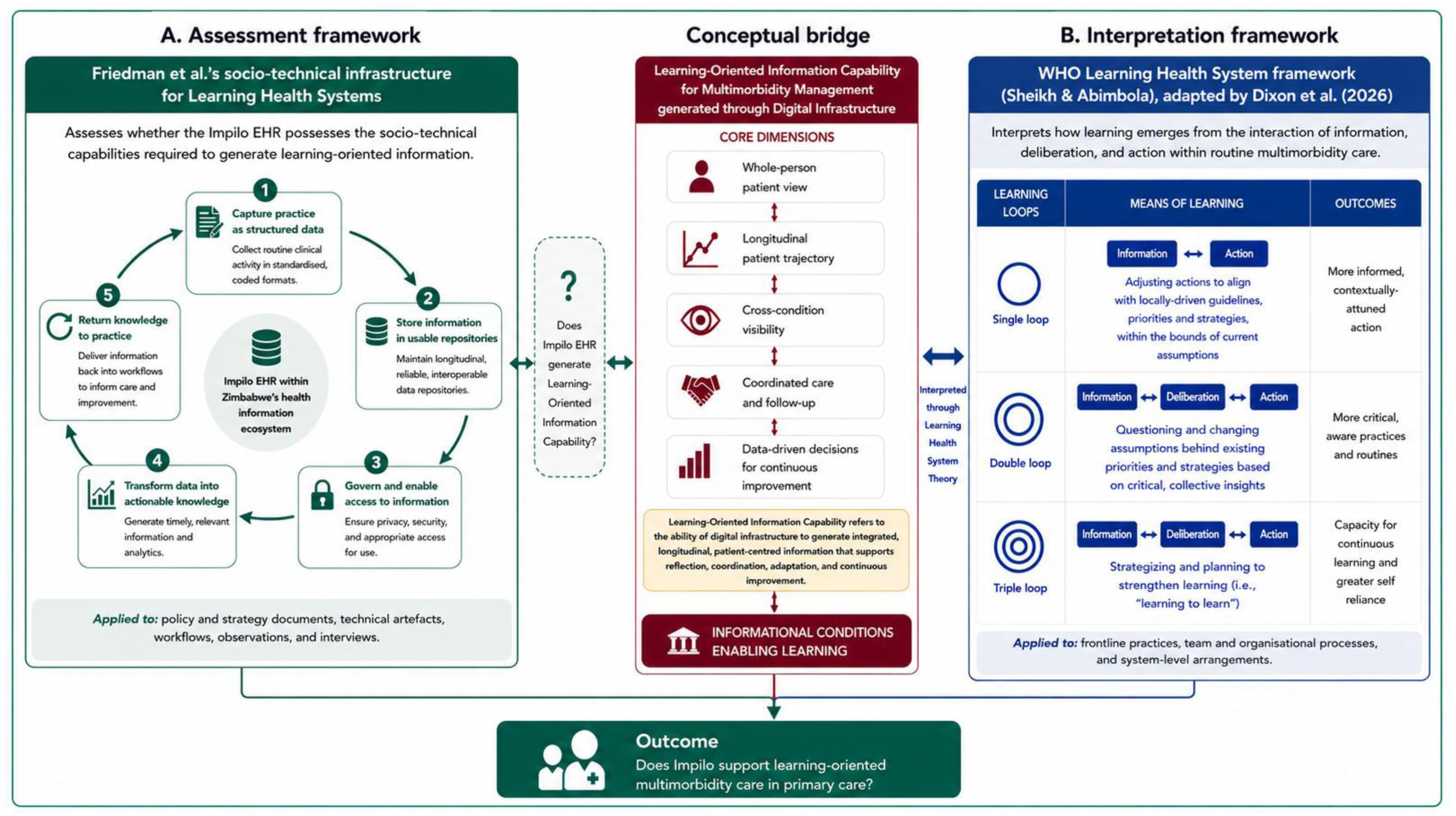
Conceptual framework for assessing multimorbidity information capability and LHS potential within Zimbabw’s digital health infrastructure. Sources: Panel A adapted from Friedman et al. (2022/2025); Panel B reproduced from Dixon et al. (2026).

In parallel, observational fieldnotes were recorded on how care processes unfolded in practice, including the use of paper tools, informal coordination, and routine problem-solving by staff. For this article, analysis focused on how these care pathways and associated learning activities were mediated by EHR, with findings triangulated against documentary and interview data.

**Fig 4.**
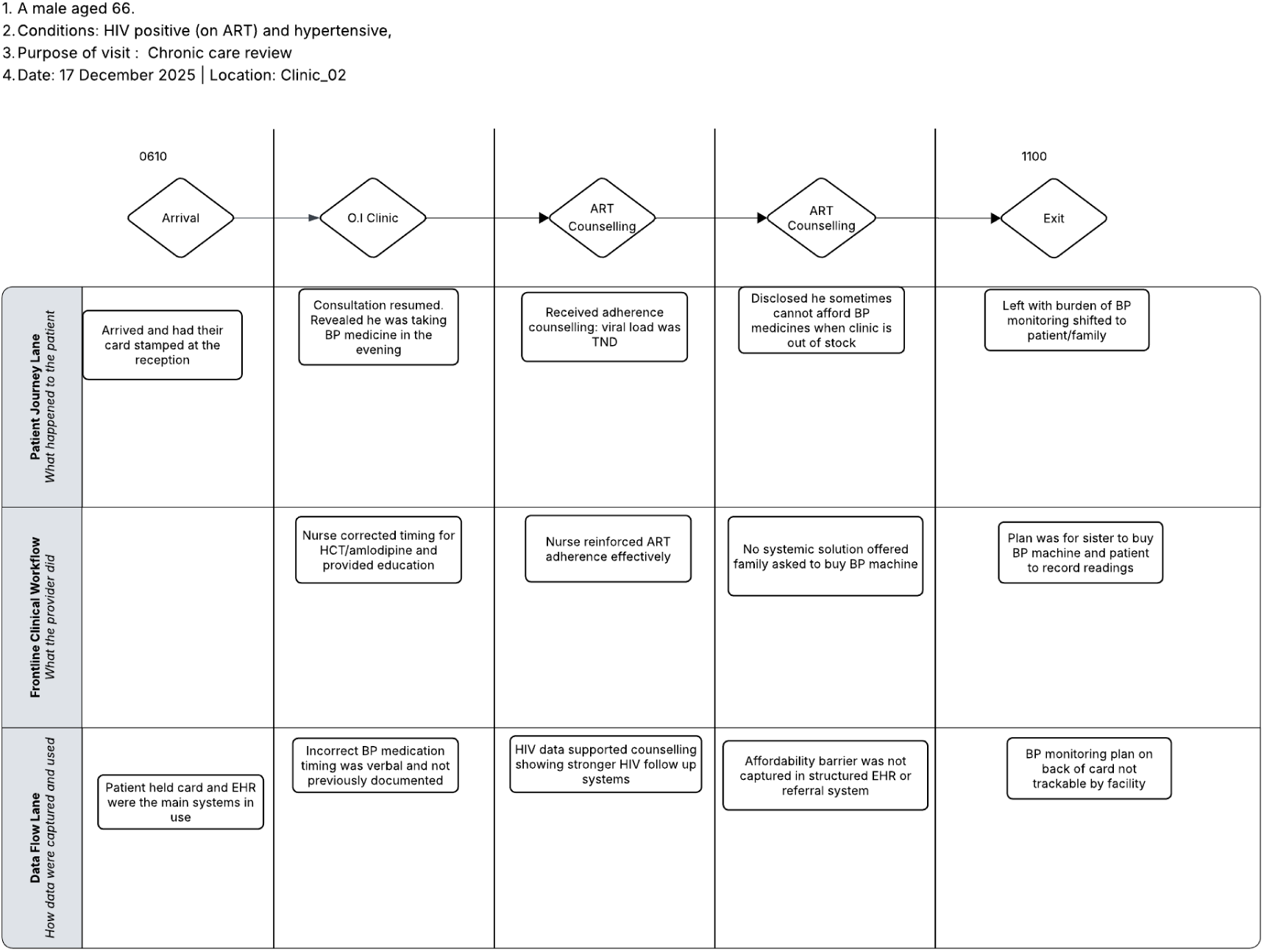
Example patient journey map illustrating interactions between patients, health workers, paper-based records, and electronic systems during routine HIV and hypertension care. (see S1 Figure for an example annotated patient journey map)

### 2.4.3 Semi-structured interviews

We conducted semi-structured interviews to examine how the Impilo EHR was designed, governed, and used in relation to multimorbidity across different levels of the health system (see table 1 for Interviewee characteristics summary). Participants were purposively sampled across system governance, programme management, technical coordination, and frontline service delivery. In total, 19 health system actors were interviewed. Drawing on methodological experience from the previous KnowM study (2), interviews were used to elicit role-specific accounts of how digital health infrastructure supported—or constrained—information use, coordination, and learning in routine care.

This multi-actor sampling strategy enabled examination of how learning, decision-making, and digital health practices were distributed across different levels of the health system (38). National and above-site actors were primarily based in Harare, with additional representation linked to Bulawayo, while facility-based interviews were conducted across clinics in Bulawayo and Chitungwiza. Written informed consent was obtained prior to participation.

Interview topic guides were structured around participants’ roles within the health system and covered processes for recording, accessing, and sharing HIV and hypertension information; use of digital and paper-based workflows across programmes and services; and routine practices related to reporting, analytics, and decision-support features. Interviews were conducted by members of the study team in private settings at participants’ workplaces or remotely via secure virtual platforms, depending on feasibility and participant preference. Interviews were audio-recorded with permission, transcribed verbatim, and anonymised. Interview duration varied according to participant role and availability, and scheduling was arranged to minimise disruption to routine service delivery.

### 2.6 Data Analysis

Documentary data were analysed using matrix-based framework analysis(*38,39*). We read all included documents in full and systematically indexed their content against the five domains. Evidence was charted into an Excel-based analytic matrix, with documents treated as analytic cases and the five domains as categories for comparison. For each document-domain combination, we recorded: (a) where relevant evidence appeared within the document (section and page reference); (b) a concise descriptive summary of how the domain was addressed; and (c) analytic memos reflecting implications for integrated, person-centred multimorbidity care. To enhance transparency and analytic consistency, descriptive extraction was distinguished from interpretive commentary, with interpretations recorded separately through analytic memos and discussed iteratively within the research team.

To support cross-document comparison and pattern recognition, we applied a simple three-point heuristic scale (0 = absent; 1 = mentioned; 2 = operationalised) to each document-domain combination. These readiness scores were assigned through an iterative analytic process led by the first author, with a purposively selected subset of documents independently reviewed by a senior anthropologist and health systems researcher (JD). Discrepancies were discussed iteratively to reach analytic consensus, consistent with recommended approaches for enhancing credibility, dependability, and analytic trustworthiness in thematic analysis(*35*). This scoring was used descriptively rather than as a measure of system performance. The completed documentary matrix was retained as a primary analytic output and subsequently imported into NVivo as secondary data to support triangulation with interview and observation findings. Table 2 summarises how Friedman’s infrastructural domains were operationalised to guide coding and analysis across documentary, interview, and observational data.

**Table 2.**
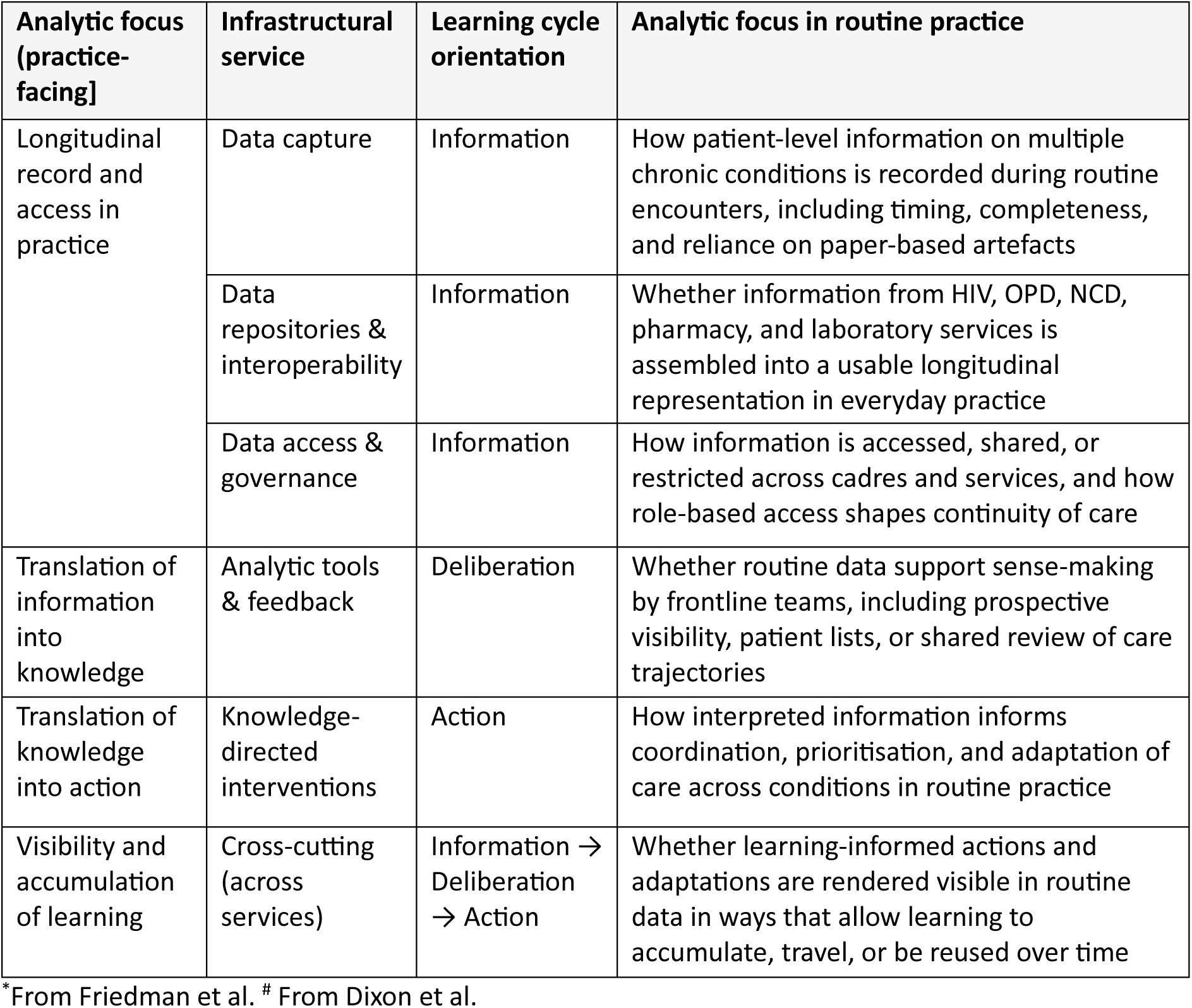
Deductive analytic framework for examining socio-technical infrastructure and learning in routine care.

| **Analytic focus (practice-facing]** | **Infrastructural service** | **Learning cycle orientation** | **Analytic focus in routine practice** |
| --- | --- | --- | --- |
| Longitudinal record and access in practice | Data capture | Information | How patient-level information on multiple chronic conditions is recorded during routine encounters, including timing, completeness, and reliance on paper-based artefacts |
|  | Data repositories & interoperability | Information | Whether information from HIV, OPD, NCD, pharmacy, and laboratory services is assembled into a usable longitudinal representation in everyday practice |
|  | Data access & governance | Information | How information is accessed, shared, or restricted across cadres and services, and how role-based access shapes continuity of care |
| Translation of information into knowledge | Analytic tools & feedback | Deliberation | Whether routine data support sense-making by frontline teams, including prospective visibility, patient lists, or shared review of care trajectories |
| Translation of knowledge into action | Knowledge-directed interventions | Action | How interpreted information informs coordination, prioritisation, and adaptation of care across conditions in routine practice |
| Visibility and accumulation of learning | Cross-cutting (across services) | Information → Deliberation → Action | Whether learning-informed actions and adaptations are rendered visible in routine data in ways that allow learning to accumulate, travel, or be reused over time |

\*From Friedman et al. # From Dixon et al.

Audio-recorded interviews were transferred to a secure shared OneDrive, transcribed verbatim, translated into English where necessary, and imported into NVivo 15 for analysis. Field notes recorded on encrypted devices were used to contextualise interview findings. An initial deductive coding framework, informed by the documentary review, focused on how information systems were enacted in practice, including longitudinal patient records, information access and exchange, and the visibility of patient care trajectories. Coding explored how Impilo and paper-based systems supported or constrained multimorbidity care, how information moved across services and cadres, and how routine documentation practices shaped care visibility. Documentary review matrices were imported into NVivo as secondary data to facilitate triangulation and interpretation of findings against identified infrastructural strengths and gaps.

To examine how these infrastructural conditions translated into learning in practice, findings from the socio-technical analysis were interpreted through the concept of learning-oriented information capability, which served as the analytical bridge between the Friedman framework and the LHS framework proposed by Abimbola and Sheikh(*17*). This enabled assessment of whether existing socio-technical arrangements generated the forms of information required to support learning across actors, encounters, and organisational levels. The LHS framework was then applied analytically rather than as a coding structure to interpret observed interactions between information, deliberation, and action in routine care, including how data were interpreted by frontline actors, how this interpretation informed action, and whether learning accumulated over time. Rather than assessing the presence or absence of a functioning LHS, the analysis focused on describing how learning-related activities were organised in practice and where learning processes became fragmented.

Alongside deductive analysis(*39*), inductive coding was undertaken to capture themes that were not fully specified in the initial framework. These included informal workarounds, experiential and reflective learning by frontline staff, reliance on patient-held artefacts, and organisational or socio-cultural factors influencing system use. Inductive codes were iteratively reviewed and examined in relation to deductive categories and learning processes, enabling refinement of interpretations while preserving a coherent analytic structure.

### 2.7 Ethics

The study was approved by the Medical Research Council of Zimbabwe ethics clearance MRCZ/A/3315. Institutional Review Board of the London School of Hygiene and Tropical Medicine, LSHTM Ethics Ref: 32628. Informed written consent was obtained for all subjects interviewed or observed in accordance with the research protocol. Data anonymisation, and confidentiality followed LSHTM guidance. Written informed consent was obtained from all participants prior to interview or observation

## 3. Results

### 3.1 Documentary Review

#### 3.1.1 Cross-domain distribution of LHS features

Documentary findings reveal uneven and largely partial specification of socio-technical infrastructure elements required to support an multimorbidity-oriented LHS across national policy, governance, programme, and evaluative documents. Across the thirteen documents (see S2 Table for documents included in documentary review), socio-technical readiness remained uneven and largely partial across domains (Table 3).

**Table 3:** Distribution of readiness scores by socio-technical domain (n = 13 documents)

| Socio-technical infrastructure domain | Readiness score = 0 (absent) | Readiness score = 1 (partially specified) | Readiness score = 2 (fully operationalised) |
| --- | --- | --- | --- |
| Data capture | 3 | 7 | 0 |
| Data repositories and interoperability | 2 | 8 | 0 |
| Access and governance | 1 | 9 | 0 |
| Analytics and feedback | 1 | 9 | 0 |
| Knowledge-directed interventions | 5 | 5 | 0 |

#### 3.1.2 Specification of Frontline System User Involvement

Across the documentary corpus, the involvement of frontline system users in the design, adaptation, or iterative refinement of digital health infrastructure was weakly articulated, even at the level of stated intent. While several documents acknowledged the importance of data use, integration, or learning in principle, few specified how frontline users were expected to participate in shaping system functionalities or adapting infrastructure in routine practice. No document described fully operationalised arrangements for user involvement across all socio-technical domains, with infrastructure most partially specified rather than embedded in everyday workflows.

This pattern was most evident in the domains of data capture and access and governance, where documents primarily positioned frontline workers as implementers of predefined documentation, reporting, and oversight arrangements. In contrast, knowledge-directed interventions were least consistently addressed, with several documents making no reference to mechanisms through which users might shape decision support or learning processes. References to analytics and feedback, as well as data repositories and interoperability, were predominantly framed in relation to institutional reporting or integration objectives, with limited articulation of routine, user-oriented feedback, reflection, or system adaptation. Notably, more explicit accounts of frontline user involvement were largely confined to context-specific research studies rather than embedded within national policy or governance documents.

### 3.2 Patient journeys

We observed 23 patient journeys involving adults receiving care for one or more long-term conditions, including HIV, hypertension, diabetes, cancer, and opportunistic infections. Participant characteristics are presented in S4 Table. Most observed journeys (17/23; 74%) involved multimorbidity, most commonly HIV and hypertension. Patient journey mapping provided formative empirical insight into how patients and healthcare providers navigated care within and across visits, enabling us to “see” and understand the patient experience as a basis for identifying opportunities for health system improvement(36).

As shown in Table 4, patients presented with one to three clinically relevant ongoing conditions, with nearly two-thirds (65%) having two conditions and a further 9% having three. Care commonly involved movement across multiple service points, with 83% of journeys traversing two or more services and almost half involving three or more. Patients encountered between zero and seven queues during a single visit and observed visit durations ranged from 17 minutes to approximately eight hours. Although the EHR was accessed during most journeys (83%), all journeys relied on paper records at some point in the care process. Four journeys involved a caregiver or treatment supporter. Collectively, these observations illustrate the complexity of routine multimorbidity care and the continued coexistence of electronic and paper-based information systems, requiring patients and providers to navigate fragmented care pathways across multiple services.

**Table 4:** Quantitative summary of observed patient journeys (n = 23)

| Descriptive characteristic* | Observed summary |
| --- | --- |
| Sex | 16 female (70%); 7 male (30%) |
| Age (years) | Median 49; range 37-76 |
| Number of clinically relevant ongoing conditions per patient | 1-3 |
| Patients with one condition | 6 (26%) |
| Patients with two conditions | 15 (65%) |
| Patients with three conditions | 2 (9%) |
| Service entry points used | OI clinic; OPD/general consultation; Family Health Services/chronic clinic; pharmacy; VIAC |
| Journeys involving ≥2 service points | 19 (83%) |
| Journeys involving ≥3 service points | 11 (48%) |
| Number of queues joined per journey | Range 0-7 |
| Duration of observed visits | 17 minutes to ~8 hours |
| Journeys involving a caregiver or treatment supporter | 4 (17%) |
| Journeys where the EHR was accessed at any point | 19 (83%) |
| Journeys relying on paper records at point of care | 23 (100%) |

\*All statistics are descriptive summaries of directly observed patient journeys; no inference to population-level prevalence is intended. Conditions were counted clinically where they were documented as diagnosed, actively treated, repeatedly monitored, investigated, or influential in care decisions during the observed journey.

### 3.3 Recording, Organising, and Using Information for care and learning

Facility-based observations extended the patient journey findings by elucidating how providers handled patient information. How they recorded, organised, and circulated it within routine service workflows, particularly where multiple services and condition-specific registers intersected. These observations made visible how digital and paper-based records were generated and distributed across departments from the outset. Interviews with fourteen system actors across national and facility levels further illuminated how these capture, storage, utilization and coordination practices were understood, navigated, and rationalised within the broader health system, providing explanatory whole system depth for the patterns observed in routine care. The five domains of Friedman’s socio-technical domains informed these observations.

#### 3.3.1 Capturing practice as Data: Point-of-care record capture and retrieval

The entry point into any LHS is data capture which is the first of the five Friedman’s socio-technical domains used in this research.

##### Tools and instruments for capturing performance

Across both learning sites, performance was recorded through a combination of paper artefacts (patient-held cards and facility registers) and the Impilo EHR as patients navigated multiple service delivery points (Table 4; S4 Table). No single instrument captured patient information across conditions or over time. For example, patients receiving hypertension care through Family Health Services were managed primarily using paper-based records (e.g. STM-01, STM-02 and STM-06), whereas patients entering HIV or OI services were first registered in the Impilo EHR before additional information was recorded in paper-based HIV booklets and clinic registers (e.g. NKT-09, NKT-10 and STM-07). Among patients with multimorbidity, information was commonly recorded across multiple paper and electronic systems during the same visit. For example, one patient with both HIV and hypertension was first registered in the EHR before being referred to OPD for hypertension management, where additional clinical information was documented separately in paper records (NKT-11). Similar patterns were observed across the patient journeys, with information distributed across multiple recording tools rather than captured within a single longitudinal patient record.

The chronic care module in Impilo, intended to capture hypertension information, was not linked to reception (the universal patient entry point) and had to be accessed separately. It did not support navigation across chronic conditions or provide a unified longitudinal view of patients across HIV and NCD modules. A senior nurse described the intended functionality:

> “We are looking forward to a vision where when I enter the patient’s name, the chronic care module should be there… so that I can navigate and check whether that patient is taking their medications well, when is their ART visit, when is their visit for NCDs. So at the moment… we can’t see that.” (HCW_01)

National stakeholders similarly described the original intention for Impilo as a longitudinal patient-centred system. One participant involved in its conceptualisation explained that it was intended to:

> “There was an existing solution… not necessarily a patient-centric system… then the discussion was around making it… a holistic patient management and decision support system”. (Decisionmaker_05)

Observations at Clinic 1 showed that hypertension screening, assessment and counselling were routinely documented but recording focused on prescribed data fields. Detailed counselling, clinical reasoning, escalation decisions and coordination across services were not routinely captured.

##### Timing and fidelity of performance capture

The timing of documentation further shaped how patient management was recorded. Interviews and observations showed that electronic documentation was frequently separated from clinical care, with data entered after consultations and often from different physical locations. One healthcare worker explained:

> “You have to go that side to enter the client… because we don’t have access to Wi-Fi here… Somebody has to move from here to go and enter them, or we ask someone to do us a favour to enter them, but they’ve got other duties.” (HCW_02)

Observations confirmed that electronic entry was commonly sequenced after patient consultations. At Clinic 2, high patient throughput and queue-based workflows prioritised patient flow, with documentation occurring in parallel or retrospectively. Patient journeys similarly showed that information was sometimes entered after clinical decisions had already been made, resulting in delayed or incomplete electronic records.

##### Capturing multimorbidity complexity

Across both sites, providers described relying on patient-held records and verbal exchange to identify coexisting conditions. During one consultation, a patient receiving long-term HIV care mentioned elevated blood pressure only as the consultation was concluding. The attending nurse remarked:

> ” Because the EHR, the electronic records only shows that this patient has a chronic disease when you open the consultation module, but when you open the HIV module, it’s not… No. It’s even invisible there, so you have to go back out… and back through”. (HCW_03)

Observations similarly showed that recognition of multimorbidity frequently depended on patient disclosure or patient-held artefacts rather than integrated electronic records. Patient journeys consistently demonstrated that information about coexisting conditions was distributed across paper records, separate electronic modules and repeated clinical encounters. Previous observations, treatment decisions and follow-up plans were not routinely visible during subsequent consultations. Overall, routine documentation reliably captured programme-specific observations and measurements but less consistently recorded coordination, clinical reasoning and interactions across multiple chronic conditions.

#### 3.3.2 Storage of information in repositories

Beyond the point of care, patient information was not stored as a single longitudinal record. OPD, HIV/OI, chronic care, pharmacy and laboratory services each generated separate records, with no unified repository representing patient care across conditions. Senior EHR technical leadership explained that this architecture reflected the system’s original deployment model:

> ““ Each facility that was receiving the solution was receiving a physical server… it was implementing in a distributed approach because of the issues of power and connectivity within the country.” (TechPartner_03_Har)

Consequently, patient information remained primarily facility-based, with central visibility dependent on manual extraction and synchronisation rather than routine real-time access. At facility level, this was experienced as fragmented information across disease modules. As one facility lead explained,

> “For the electronic health records, therès no integration. The chronic disease, they have their own module, the HIV, they have their own module, so it’s up to the clinicians to know that this patient is also this chronic disease.” (HCW_03)

The absence of an integrated repository also limited visibility of multimorbidity at population level. Another participant noted:

> “ Actually, chronic care has gotten WORSE since going paperless… they were following up the patients, but since the EHR version barely exists, they’re not and forgetting to follow up“. (HCW_05)

Observations and patient journeys similarly showed that paper records were not routinely incorporated into electronic repositories, and information recorded in separate modules was not automatically assembled into a longitudinal patient record. As another clinician observed:

> “ It’s ‘hypertension, hypertension, hypertension’… wère not seeing the diabetes, therès a gap…” (HCW_05)

#### 3.3.3 Governed access to data for decision making

Documentary review showed well-defined governance arrangements, including role-based access control, documented authorisation procedures, audit logging, secure authentication and regular access reviews. These arrangements primarily emphasised security, accountability and managerial oversight. At facility level, however, governed access was experienced mainly through reporting responsibilities rather than cross-condition clinical visibility.

Governance was evident in routine supervision of data quality and completeness. A Sister in Charge described reviewing tablet entries to ensure:

> “Every field has been filled”

and mentoring staff where chronic diagnoses had been omitted, noting that

> “ Those patients do not appear.” (HCW_01)

Governance was therefore embedded not only in policy but in routine supervisory practice. However, access to information for clinical decision-making remained strongly role-bound and sequential. Observations showed care organised as task-specific activities—registration, consultation, dispensing, and data entry—each tied to designated users and devices. Connectivity constraints further shaped access. As one healthcare worker explained:

> “You have to go that side to enter the client… because we don’t have access to that module here” (HCW_02)

Patient journeys similarly showed that clinically relevant information recorded at one point of care was not consistently visible across subsequent service points during the same visit. Consequently, governed access supported accountability and reporting but was less aligned with the information needs of integrated clinical decision-making.

#### 3.3.4 Data to knowledge: limited patient level prospective sense-making

Across both learning sites, information systems did not routinely generate prospective outputs such as integrated patient line lists, synchronised appointments or shared reminders for patients with multimorbidity. Knowledge about patient status therefore emerged primarily during clinical encounters rather than from accumulated longitudinal data. A senior facility lead explained:

> “ A line list… means planning for proper care not haphazardly… need glucometer sticks, BP… to deliver the supermarket approach that patients expect.” (HCW_05)

While the EHR generated programme-specific registers, these did not routinely provide integrated views of patients with multiple chronic conditions. Recognition of multimorbidity therefore depended on patient disclosure, clinician recall or manual cross-checking. Observations and patient journeys similarly showed that longitudinal patient information was reconstructed through patient-held records, clinical hand-offs and retrospective review rather than retrieved from a single electronic record.

At facility level, aggregate reports were routinely reviewed during monthly programme meetings, particularly for HIV services. EHR outputs and paper registers were reconciled, and performance against programme targets discussed. These deliberations focused primarily on programme-specific indicators, with limited attention to patients with multiple chronic conditions. Similar programme-specific review structures were observed at district and city level. Reflecting on this organisational context, one national stakeholder explained:

> “ What then came through over the years was verticalization… taking one area of knowledge, one disease condition, then you say, strictly stick to this because that’s what is funded.” (Decisionmaker_02)

Knowledge generation therefore depended largely on individual initiative and retrospective review. As one clinician described:

> “When you finish your job, run your own report and see what you have done… then you start to see the missing pieces.” (HCW_06).

Overall, patient-level knowledge remained largely encounter-based, while routine system outputs primarily supported programme-specific reporting.

#### 3.3.5 Knowledge to performance: adaptive and experiential action

Knowledge generated during patient encounters routinely informed clinical action. Across both learning sites, providers adapted care through counselling, referral, treatment adjustment and prioritisation based on information assembled during consultations. Observations showed that recognised clinical risks were managed according to established protocols. For example, elevated blood pressure prompted immediate treatment and repeat measurement before further clinical decisions were made.

Interview data similarly showed that clinicians frequently compensated for limited system visibility by actively seeking additional information. As one clinician explained:

> “ If you have a functioning chronic register… patient should feel their nurse is there… but it’s not if you have to ask all the basic information again”. (HCW_05)

Patient journeys reinforced these observations. Clinical actions were commonly informed by information reconstructed from patient-held records, questioning and consultation findings rather than from integrated electronic records. Although care was adapted during individual encounters, these adaptations were not consistently visible during subsequent visits or across service points. Knowledge use also depended on individual initiative. As one clinician described:

> “When you finish your job, run your own report and see what you have done… then you start to see the missing pieces.” (HCW_06)

Overall, knowledge informed routine clinical action, but adaptive practices remained largely encounter-based and individually enacted rather than consistently embedded within longitudinal electronic records.

## 4. Discussion

Amid rising multimorbidity globally, EHRs are increasingly positioned as critical infrastructure for supporting integrated, person-centred care and enabling health systems to learn from routine practice (40). This expectation is particularly important in resource-constrained settings, where routine data represent one of the few scalable resources available for responding to growing clinical complexity. Using Zimbabw’s national Impilo EHR as a case study, we examined whether existing digital health infrastructure supports the conditions required for learning-oriented multimorbidity care. Rather than evaluating system performance alone, which has already been done (15,19) this study explored how socio-technical arrangements around Impilo EHR design, deployment and utilization shape the emergence of learning in routine care(18). We identified a disconnect between policy aspirations for integrated, patient-centred care and the information capabilities available to support such care in practice. These findings suggest that the challenge is not simply the presence or absence of digital infrastructure, but the extent to which it enables learning across patients, providers, services, and organisational levels.

The sections that follow examine this finding in greater depth. We first consider the predominance of single-loop learning in routine care, before exploring learning-oriented information capability as a bridge between digital infrastructure and learning processes. We then examine how frontline providers compensate for these limitations through workarounds and finally consider co-production as a mechanism for strengthening learning-oriented multimorbidity care.

### 4.1 Learning in routine care occurs, but is predominantly single loop

Across the selected primary care settings with relatively advanced EHR use, a consistent pattern emerged: frontline providers continuously used individual clinical guidelines to interpret patient information, coordinate care across services, and adapt clinical decisions within existing organisational and resource constraints. In this sense, learning was clearly occurring. However, that learning remains largely embedded within individual encounters and local problem-solving rather than being systematically captured, shared, or accumulated across the wider health system. (2). Patients frequently acted as the primary carriers of continuity through disclosure, treatment histories, and paper-based records, enabling providers to reconstruct fragmented care trajectories. While these practices supported ongoing care, they also revealed the extent to which learning depended on individual actors rather than institutionalised information processes. As a result, opportunities for cross-condition learning and adaptation were rarely translated into organisational memory or system-wide improvement.

Viewed through our adapted LHS framework, these findings are most consistent with single-loop learning, whereby providers adjust actions within existing routines but rarely influence the underlying structures, assumptions, or pathways through which care is organised (17). Although learning emerged through interactions between information, deliberation, and action(11), it was not routinely converted into cumulative, shareable, and actionable knowledge capable of informing broader adaptation. This finding echoes wider LHS scholarship showing that the generation of data alone does not produce learning(16); rather, learning depends on socio-technical arrangements that enable knowledge to circulate across individuals, services, and organisational levels. As Friedman et al., argue, the challenge, therefore, was not an absence of learning, but the limited capacity of existing information infrastructure to transform local learning into a collective system resource.(18).

### 4.2 Learning-oriented information capability: the missing between socio-technical infrastructure and learning?

A central finding of this study is that although policy and system design documentation envisioned integrated, patient-centred records within the Impilo EHR (19), the system was not able to generate the forms of information required for enabling double loop learning for multimorbidity care. These include whole-patient summaries, synchronised patient line lists, prospective review schedules, and other tools that support care across conditions rather than reproducing disease-specific workflows (32). In this sense, Impilo EHR demonstrates constrained learning-oriented information capability. We define learning-oriented information capability as the ability of health information infrastructure to generate, organise, and make visible information that supports collective interpretation, coordination, feedback, and improvement across time and organisational levels. This definition is derived from well-respected scholarship on LHS and quality improvement that that emphasises the importance of information infrastructures in supporting collective sensemaking and adaptation (41–44).

These findings suggest that learning-oriented information capability may provide a useful analytical bridge between socio-technical infrastructure and LHS processes. While socio-technical frameworks describe the infrastructures required to generate and manage information (45), and LHS frameworks explain how learning emerges through interactions between information, deliberation, and action (16,17), the informational conditions linking these domains have received less explicit attention. Our findings indicate that the ability to generate whole-person patient views, longitudinal trajectories, cross-condition visibility, coordinated follow-up, and practice-facing information may represent important preconditions for learning-oriented multimorbidity care.

### 4.3 Workarounds as frontline health workers sustaining fragmented care architectures

There is growing recognition that effective multimorbidity care requires not only service integration but also person-centred approaches that respond to the needs of individuals living with multiple chronic conditions. Programmes such as INTE-Africa have demonstrated the feasibility and benefits of integrating HIV and NCD services, contributing to broader policy and programmatic efforts to move beyond disease-specific models of care(6). However, our findings suggest that less attention has been paid to the routine information capabilities required to support integrated, person-centred care in everyday clinical practice.

The patient journeys examined in this study revealed that frontline providers were already attempting to deliver integrated, person-centred care. Clinicians routinely constructed whole-person understandings of patients by eliciting histories across conditions, reconciling information from multiple sources, coordinating across services, and adapting care to evolving patient circumstances. Yet these activities were rarely supported by the underlying information infrastructure. Instead, providers relied on paper artefacts, informal communication, and retrospective review of their own documentation to maintain continuity of care. In effect, frontline providers became the primary mechanism through which fragmented information, services, and care pathways were brought together.

These efforts gave rise to workarounds through which providers sought to compensate for the limited learning-oriented information capability described above. Workarounds therefore function as diagnostic signals of socio-technical misalignment. They reveal a system in which the aspiration for integrated, person-centred multimorbidity care increasingly exceeds the information capabilities available to support it. Similar patterns have been observed in other resource-constrained settings, where EHR systems expose or amplify workflow misalignments and require users to compensate through informal practices rather than benefiting from redesigned, learning-oriented processes (47,48).

From the perspective of Abimbola and Sheikh’s LHS framework, this represents a missed opportunity for double-loop learning(17). All the ingredients were there. Frontline providers routinely generated insights from multimorbidity encounters, care coordination challenges, and fragmented patient journeys. These experiences provided a rich source of information about the limitations of existing disease-specific approaches to care and the practical requirements of integrated, person-centred service delivery. However, such insights were rarely translated into forms of information that could be accumulated, shared, and deliberated upon across organisational levels, something critical for single loop learning to evolve into double loop learning. A missed opportunity for the health system to use frontline experience to question and adapt the assumptions, approaches and verticalized arrangements currently underpinning multimorbidity care.

### 4.4 Reconnecting learning and system design: co-production as a mechanism

These findings have important implications for the development of Learning Health Systems in resource-constrained settings at a time when the need for adaptive approaches to multimorbidity care has never been greater. Across many LMICs, rising burdens of chronic disease alongside HIV are coinciding with growing recognition that health systems must become more capable of learning from routine practice and adapting to changing needs(2). Recent reductions in external funding have further reinforced the importance of locally owned, sustainable approaches to health system strengthening, increasing interest among governments and Ministries of Health in learning, adaptation, and self-reliance as core system capabilities(49).

Viewed from this perspective, the gap identified in this study represents a significant opportunity. The principal constraint was not the absence of data, digital infrastructure, or frontline learning capacity. Rather, it was the limited ability of existing socio-technical arrangements to transform routine experience into information that could support collective reflection, coordinated action, and system-level adaptation. Importantly, Zimbabwe is well positioned to address this challenge. Unlike many externally developed digital health platforms, Impilo is nationally owned, locally developed, and continues to evolve. The ongoing development of its chronic care module therefore provides a rare opportunity to embed learning-oriented information capability within the architecture of the system itself (26).

Evidence from digital health and implementation science shows that involving frontline actors in defining these elements improves alignment with practice and supports cumulative learning(50,51). Reconnecting frontline learning to system design through co-production therefore represents a promising pathway for addressing the socio-technical misalignments identified in this study. By involving users throughout the design, development, and iterative refinement process, co-production can help ensure that digital systems generate the longitudinal patient views, coordinated workflows, cross-condition visibility, and practice-facing information required to support learning-oriented multimorbidity care(52,53). To synthesise these findings, Table 2 summarises the principal socio-technical constraints identified in this study and the corresponding co-production-enabled responses relevant to strengthening longitudinal, learning-oriented multimorbidity care.

**Table 5:** How co-production reorients digital system design for multimorbidity care.

| Dominant design orientation in the existing system | How this manifested in routine multimorbidity care | What co-production reorients |
| --- | --- | --- |
| Disease-specific programme architectures | Patients navigated parallel HIV, hypertension, and other chronic pathways with fragmented records and duplicated encounters | Design shifts toward a unified chronic care “space” organised around whole-patient continuity |
| Reporting-oriented data structures | Data supported programme accountability more than longitudinal clinical care | Frontline actors prioritised practice-facing tools such as shared review lists, longitudinal summaries, and coordinated follow-up |
| Top-down system configuration | Frontline staff relied on workarounds, memory, and informal coordination to sustain care | Experiential frontline knowledge becomes a core input into workflow and system design |
| Separate disease scheduling systems | Patients attended multiple unsynchronised visits despite stable chronic conditions | Follow-up logic redesigned around coordinated multimorbidity care pathways |
| Modular visibility boundaries | Providers could not easily see all active chronic conditions, medicines, or referrals together | Co-production prioritised longitudinal multimorbidity visibility across services and encounters |
| Retrospective and duplicated data capture | Clinicians balanced patient care with parallel paper and EHR documentation burdens | Workflows redesigned around capture-once, reuse-many-times principles aligned with routine care |
| Static development processes | Persistent mismatch between evolving care realities and digital tools | Iterative refinement and frontline validation embedded into system evolution |
| Programme-centred reporting logic | Patients became represented as separate diseases rather than whole persons | Analytics redesigned to support person-level multimorbidity visibility and learning |
| Separation of technical design from everyday care realities | Learning remained localised, informal, and difficult to translate into system adaptation | Co-production reconnects frontline learning with ongoing system redesign and adaptation |

## 5. Conclusion

This study showed that although Zimbabw’s national EHR was conceived as a patient-centred system, it currently lacks the capacity to generate the integrated, longitudinal, and cross-condition information required to support learning-oriented multimorbidity care. The principal constraint lies not in data availability, digital infrastructure, or frontline learning capacity, but in the system’s limited ability to transform routine clinical information into a resource for collective reflection, coordinated action, and adaptation. As a result, learning remains largely encounter-bound, with limited capacity to inform broader system change. For African countries investing in national EHRs, the challenge is not simply to digitise care, but to design systems that generate the informational foundations for whole-person care, continuous learning, and adaptation. Without such alignment, digital systems risk reinforcing existing silos; with it, they may become important enablers of integrated, adaptive care in contexts of rising multimorbidity and constrained resources. These findings suggest that information capability may represent a critical bridge between socio-technical infrastructure and Learning Health System processes. Co-production offers a practical pathway for achieving this by connecting frontline experience to system design and governance.

## 6. Strengths and Limitations

Strengths of this study include purposive selection of analytically rich sites, prolonged engagement in the field, and triangulation across observations, interviews, and documentary sources. However, the analysis was limited to two learning sites and on Impilo as the dominant national EHR. As such, findings are analytically rather than statistically, generalisable and may not capture learning dynamics in parallel or locally adapted systems. The study was diagnostic rather than evaluative, focusing on the conditions that enable or constrain learning rather than on system performance. Future research should assess how co-produced EHR modifications, including longitudinal and analytic artefacts, support collective sensemaking and system-level adaptation.

## Supporting information

S1 SRQR Checklist

S2 Documentary review

S3 Participant summary

S4 Patient journey

## Data Availability

Data supporting the findings of this study have been deposited with the UK Data Service ReShare repository. Access is subject to governance procedures and participant confidentiality protections.

## Author Contributions

**Conceptualization:** Efison Dhodho, Justin Dixon, Dorothea Nitsch

**Methodology:** Efison Dhodho, Justin Dixon, Dorothea Nitsch.

**Software:** Efison Dhodho, Forget Banda, Blessing Manyiyo, Robert T Gongora

**Investigation:** Efison Dhodho, Kety Choga, Fiona Mundoga, Forget Banda, Agnes Katsidzira, Trudy Mhlanga, Justin Dixon.

**Data curation:** Efison Dhodho, Kety Choga, Fiona Mundoga, Kenneth Masiye.

**Formal analysis:** Efison Dhodho, Fiona Mundoga, Justin Dixon

**Visualization:** Kenneth Masiye.

**Funding acquisition:** Justin Dixon, Efison Dhodho, Karen Webb, Theonevus T. Chinyanga, Pugie T Chimberengwa

**Resources:** Karen Webb, Theonevus T. Chinyanga, Pugie T Chimberengwa

**Project administration:** Kety Choga, Karen Webb, Theonevus T. Chinyanga, Trudy Mhlanga, Pugie T Chimberengwa

**Supervision:** Nicholas Midzi, Justin Dixon, Dorothea Nitsch.

**Validation:** Fiona Mundoga, Pugie T. Chimberengwa, Robert Tawanda Gongora, Trymore Chaurura, Nicholas Midzi, Justice Mudavanhu, Patience Mangisi, Clorata Gwanzura, Shingirirayi Tsvangirayi, Dorothea Nitsch.

**Writing - original draft:** Efison Dhodho.

**Writing - review & editing:** Kety Choga, Fiona Mundoga, Pugie T. Chimberengwa, Robert Tawanda Gongora, Trymore Chaurura, Karen Webb, Theonevus T. Chinyanga, Forget Banda, Kenneth Masiye, Nicholas Midzi, Justice Mudavanhu, Agnes Katsidzira, Blessing Manyiyo, Tsitsi Apollo, Cleopas Chimbetete, Trudy Mhlanga, Patience Mangisi, Clorata Gwanzura, Shingirirayi Tsvangirayi, Blessing Manyiyo, Memory Benjamin, Justin Dixon, and Dorothea Nitsch, Valiant Makore

All authors reviewed and approved the final manuscript.

## 7. Acknowledgements

We thank the Zimbabwe Ministry of Health and Child Care, Provincial and City Health authorities in Harare and Bulawayo Metropolitan Provinces, the University of Zimbabwe, the National University of Science and Technology, the Organisation for Public Health Interventions and Development, and colleagues from the World Health Organisation in Zimbabwe for their invaluable guidance, support and collaboration. Our sincere gratitude to all the Nketa and St Marys Clinic staff for their ready participation in the study. Their engagement and support were essential to the successful completion of this research.

## 8. Availability of data and materials (paster the link)

Data are available upon reasonable request. Anonymous survey responses will be made available on reasonable request to the authors.

## 11. Authors’ contributions

Efison Dhodho conceptualised the study, developed the methodology, conducted formal analysis, led visualisation, managed the project, and wrote the original manuscript draft. KC conducted ethnographic observations, patient journey mapping and shadowing, file management, and financial administration for the study. FM conducted ethnographic observations and patient journey mapping, supported participant recruitment and consent processes, and contributed to field coordination activities. Justin Dixon contributed to methodology development, provided supervision throughout the study, and reviewed and edited the manuscript. Dorothea provided clinical interpretation and supervision and contributed to manuscript review and editing. Professor Nicholas Midzi facilitated ethical clearance processes and reviewed study documents. Agnes supported ethnographic observations and reviewed the manuscript. All authors reviewed and approved the final manuscript.

## References

1. Gouda HN, Charlson F, Sorsdahl K, Ahmadzada S, Ferrari AJ, Erskine H, et al. Burden of non-communicable diseases in sub-Saharan Africa, 1990-2017: results from the Global Burden of Disease Study 2017. Lancet Glob Health. 2019 Oct;7(10):e1375–87. doi:10.1016/S2214-109X(19)30374-2

2. Dixon J, Dhodho E, Mundoga F, Webb K, Chimberengwa P, Mhlanga T, et al. Multimorbidity and health system priorities in Zimbabwe: A participatory ethnographic study. Haghparast Bidgoli H, editor. PLOS Glob Public Health. 2025 Apr 28;5(4):e0003643. doi:10.1371/journal.pgph.0003643

3. Fortin M, Stewart M, Poitras ME, Almirall J, Maddocks H. A Systematic Review of Prevalence Studies on Multimorbidity: Toward a More Uniform Methodology. Ann Fam Med. 2012 Mar 1;10(2):142–51. doi:10.1370/afm.1337

4. Kwarisiima D, Balzer L, Heller D, Kotwani P, Chamie G, Clark T, et al. Population-Based Assessment of Hypertension Epidemiology and Risk Factors among HIV-Positive and General Populations in Rural Uganda. Lima VD, editor. PLOS ONE. 2016 May 27;11(5):e0156309. doi:10.1371/journal.pone.0156309

5. SANOFI. Sanofi: Taking the lead in global health: Global Health Unit Impact Report [Internet]. Paris: SANOFI; 2025 [cited 2025 Feb 18]. Available from: https://ghu-impact-report.sanofi.com/article/1/?utm_source=chatgpt.com

6. Kivuyo S, Birungi J, Okebe J, Wang D, Ramaiya K, Ainan S, et al. Integrated management of HIV, diabetes, and hypertension in sub-Saharan Africa (INTE-AFRICA): a pragmatic cluster-randomised, controlled trial. The Lancet. 2023 Oct;402(10409):1241–50. doi:10.1016/S0140-6736(23)01573-8

7. Endalamaw A, Zewdie A, Wolka E, Assefa Y. A scoping review of digital health technologies in multimorbidity management: mechanisms, outcomes, challenges, and strategies. BMC Health Serv Res. 2025 Mar 15;25(1):382. doi:10.1186/s12913-025-12548-5

8. Forslund M, Mathieson K, Djibo Y, Mbindyo C, Lugangira N, Balasubramaniam P. Strengthening the evidence base on the use of digital health technologies to accelerate progress towards universal health coverage. Oxf Open Digit Health. 2024 Jan 1;2:oqae033. doi:10.1093/oodh/oqae033

9. Moucheraud C, Schwitters A, Boudreaux C, Giles D, Kilmarx PH, Ntolo N, et al. Sustainability of health information systems: a three-country qualitative study in southern Africa. BMC Health Serv Res. 2017 Dec;17(1):23. doi:10.1186/s12913-016-1971-8

10. Mandreoli F, Ferrari D, Guidetti V, Motta F, Missier P. Real-world data mining meets clinical practice: Research challenges and perspective. Front Big Data. 2022 Oct 21;5:1021621. doi:10.3389/fdata.2022.1021621

11. Witter S, Sheikh K, Schleiff M. Learning health systems in low-income and middle-income countries: exploring evidence and expert insights. BMJ Glob Health. 2022 Sep;7(Suppl 7):e008115. doi:10.1136/bmjgh-2021-008115

12. Dixon J, Dhodho E, Webb K, Chimberengwa P, Mundoga F, Choga K, et al. Multimorbidity: a core priority for learning health systems amidst vertical disease programme cuts. Health Res Policy Syst. 2026 Feb 19;24(1):20. doi:10.1186/s12961-026-01456-7

13. Angwenyi V, Aantjes C, Kajumi M, De Man J, Criel B, Bunders-Aelen J. Patients experiences of self-management and strategies for dealing with chronic conditions in rural Malawi. PLOS ONE. 2018 Jul 2;13(7):7. Located at: WOS:000437224100034. doi:10.1371/journal.pone.0199977

14. Eyowas FA, Schneider M, Alemu S, Getahun FA. Experience of living with multimorbidity and health workers perspectives on the organization of health services for people living with multiple chronic conditions in Bahir Dar, northwest Ethiopia: a qualitative study. BMC Health Serv Res. 2023 Mar 9;23(1):232. doi:10.1186/s12913-023-09250-9

15. Dumisani Mugauri H, Chimsimbe M. Electronic Health Record Systems in Limited Resource Settings: A Comprehensive Evaluation of the Impilo Platform. Acta Inform Pragensia. 2025 May 7. doi:10.18267/j.aip.265

16. Enticott J, Johnson A, Teede H. Learning health systems using data to drive healthcare improvement and impact: a systematic review. BMC Health Serv Res. 2021 Dec;21(1):200. doi:10.1186/s12913-021-06215-8

17. Abimbola Seye, Sheikh Abir. Learning Health Systems: Pathways to Progress. Flagship Report of the Alliance for Health Policy and Systems Research. 1st ed. Geneva: World Health Organization; 2021. 1 p.

18. Friedman CP, Lomotan EA, Richardson JE, Ridgeway JL. Socio-technical infrastructure for a learning health system. Learn Health Syst. 2024 Jan;8(1). doi:10.1002/lrh2.10405

19. MOHCC. Process Evaluation of the Impilo Electronic Health Record (EHR) System at Selected Health Facilities in Zimbabwe [Evaluation Report]. Harare: Ministry of Health and Child Care, Zimbabwe; 2025.

20. Mugauri HD, Chimsimbe M, Shambira G, Shamhu S, Nyamasve J, Munyanyi M, et al. A decade of designing and implementing electronic health records in Sub-Saharan Africa: a scoping review. Glob Health Action. 2025 Dec 31;18(1):2492913. doi:10.1080/16549716.2025.2492913

21. Institute of Medicine. Digital Infrastructure for the Learning Health System: The Foundation for Continuous Improvement in Health and Health Care: Workshop Series Summary [Internet]. Washington, D.C.: National Academies Press; 2011 [cited 2026 Jun 7]. Available from: https://www.nationalacademies.org/publications/12912 doi:10.17226/12912

22. Butler JM, Gibson B, Lewis L, Reiber G, Kramer H, Rupper R, et al. Patient-centered care and the electronic health record: exploring functionality and gaps. JAMIA Open. 2020 Oct;3(3):3. doi:10.1093/jamiaopen/ooaa044 PubMed PMID: 33215071; PubMed Central PMCID: PMC7660957.

23. OECD. Building people-centred digital health systems: Lessons from PaRIS [Internet]. 2026 Mar [cited 2026 Jun 7]. Available from: https://www.oecd.org/en/publications/building-people-centred-digital-health-systems_a1df0046-en.htmldoi:10.1787/a1df0046-en

24. Dhodho E, Masiye K, Banda F, Bepe T, Nyathi N, Chinyanga TT. Transforming Routine Health Data Use in LMICs through Modular, AI-Supported Automation: Insights from Zimbabwe. Oxf Open Digit Health. 2026 Jan 30;oqag003. doi:10.1093/oodh/oqag003

25. ÒBrien BC, Harris IB, Beckman TJ, Reed DA, Cook DA. Standards for Reporting Qualitative Research: A Synthesis of Recommendations. Acad Med. 2014 Sep;89(9):1245–51. doi:10.1097/ACM.0000000000000388

26. MOHCC. Impilo Electronic Health Record System. Government of Zimbabwe; 2023.

28. Palinkas LA, Horwitz SM, Green CA, Wisdom JP, Duan N, Hoagwood K. Purposeful Sampling for Qualitative Data Collection and Analysis in Mixed Method Implementation Research. Adm Policy Ment Health Ment Health Serv Res. 2015 Sep;42(5):533–44. doi:10.1007/s10488-013-0528-y

29. Dixon J, Dhodho E, Mundoga F, Webb K, Chimberengwa P, Mhlanga T, et al. Multimorbidity and health system priorities in Zimbabwe: A participatory ethnographic study [Internet]. 2024 Aug 7. doi:10.1371/journal.pgph.0003643

30. Aqil A, Lippeveld T, Hozumi D. PRISM framework: a paradigm shift for designing, strengthening and evaluating routine health information systems. Health Policy Plan. 2009 May 1;24(3):217–28. doi:10.1093/heapol/czp010

31. Sari DSK, Lipoeto NI, Bachtiar H, Catri I, Sari NK, Semiarty R. Assessing Telemedicine Demand and Viability in Indonesian Geriatric Clinics: A Comprehensive HOT FIT and Sociotechnical Analysis. Curr Aging Sci. 2025 Mar;18(1):47–58. doi:10.2174/0118746098302999240522092726

32. Butler JM, Gibson B, Lewis L, Reiber G, Kramer H, Rupper R, et al. Patient-centered care and the electronic health record: exploring functionality and gaps. JAMIA Open. 2020 Oct;3(3):360–8. doi:10.1093/jamiaopen/ooaa044 PubMed PMID: 33215071; PubMed Central PMCID: PMC7660957.

33. Pawa J, Robson J, Hull S. Building managed primary care practice networks to deliver better clinical care: a qualitative semi-structured interview study. Br J Gen Pract. 2017 Nov;67(664):664. doi:10.3399/bjgp17X692597

34. Dalglish SL, Khalid H, McMahon SA. Document analysis in health policy research: the READ approach. Health Policy Plan. 2021 Feb 16;35(10):1424–31. doi:10.1093/heapol/czaa064

35. Nowell LS, Norris JM, White DE, Moules NJ. Thematic Analysis: Striving to Meet the Trustworthiness Criteria. Int J Qual Methods. 2017 Dec 1;16(1):1609406917733847. doi:10.1177/1609406917733847

36. Trebble TM, Hansi N, Hydes T, Smith MA, Baker M. Process mapping the patient journey: an introduction. BMJ. 2010 Aug 13;341(aug13 1):c4078–c4078. doi:10.1136/bmj.c4078

37. Bitner MJ, Ostrom AL, Morgan FN. Service Blueprinting: A Practical Technique for Service Innovation. Calif Manage Rev. 2008 Apr;50(3):66–94. doi:10.2307/41166446

38. Goldsmith L. Using Framework Analysis in Applied Qualitative Research. Qual Rep. 2021 Jun 20. doi:10.46743/2160-3715/2021.5011

39. Gale NK, Heath G, Cameron E, Rashid S, Redwood S. Using the framework method for the analysis of qualitative data in multi-disciplinary health research. BMC Med Res Methodol. 2013 Sep 18;13(1):117. doi:10.1186/1471-2288-13-117

40. Odekunle FF, Odekunle RO, Shankar S. Why sub-Saharan Africa lags in electronic health record adoption and possible strategies to increase its adoption in this region. Int J Health Sci. 2017;11(4):59–64. PubMed PMID: 29085270; PubMed Central PMCID: PMC5654179.

41. Irimu G, Ogero M, Mbevi G, Agweyu A, Akech S, Julius T, et al. Approaching quality improvement at scale: a learning health system approach in Kenya. Arch Dis Child. 2018 Mar 7;archdischild-2017-314348. doi:10.1136/archdischild-2017-314348

42. Tuti T, Aluvaala J, Chelangat D, Mbevi G, Wainaina J, Mumelo L, et al. Improving in-patient neonatal data quality as a pre-requisite for monitoring and improving quality of care at scale: A multisite retrospective cohort study in Kenya. Asweto CO, editor. PLOS Glob Public Health. 2022 Oct 20;2(10):e0000673. doi:10.1371/journal.pgph.0000673

43. Ainsworth J, Buchan I. Combining Health Data Uses to Ignite Health System Learning. Methods Inf Med. 2015;54(06):479–87. doi:10.3414/ME15-01-0064

44. Gilson L, Barasa E, Brady L, Kagwanja N, Nxumalo N, Nzinga J, et al. Collective sensemaking for action: researchers and decision makers working collaboratively to strengthen health systems. BMJ. 2021 Feb 15;m4650. doi:10.1136/bmj.m4650

45. Sittig DF, Singh H. A new sociotechnical model for studying health information technology in complex adaptive healthcare systems. Qual Saf Health Care. 2010 Oct 1;19(Suppl 3):i68–74. doi:10.1136/qshc.2010.042085

46. Hardie T, Horton T, Thornton-Lee N, Home J, Pereira P. Developing learning health systems in the UK: Priorities for action [Internet]. The Health Foundation; 2022 Sep [cited 2026 Jan 30]. Available from: https://www.health.org.uk/publications/reports/developing-learning-health-systems-in-the-uk-priorities-for-action doi:10.37829/HF-2022-I06

47. Greenhalgh T, Potts HWW, Wong G, Bark P, Swinglehurst D. Tensions and Paradoxes in Electronic Patient Record Research: A Systematic Literature Review Using the Meta-narrative Method. Milbank Q. 2009 Dec;87(4):729–88. doi:10.1111/j.1468-0009.2009.00578.x

48. Zharima C, Griffiths F, Goudge J. Exploring the barriers and facilitators to implementing electronic health records in a middle-income country: a qualitative study from South Africa. Front Digit Health. 2023;5:1207602. doi:10.3389/fdgth.2023.1207602 PubMed PMID: 37600481; PubMed Central PMCID: PMC10437058.

49. Dixon J, Dhodho E, Webb K, Chimberengwa P, Mundoga F, Choga K, et al. Multimorbidity: a core priority for learning health systems amidst vertical disease programme cuts. Health Res Policy Syst. 2026 Feb 19;24(1):20. doi:10.1186/s12961-026-01456-7

50. Gannon H, Chimhuya S, Chimhini G, Neal SR, Shaw LP, Crehan C, et al. Electronic application to improve management of infections in low-income neonatal units: pilot implementation of the NeoTree beta app in a public sector hospital in Zimbabwe. BMJ Open Qual. 2021 Jan;10(1):1. doi:10.1136/bmjoq-2020-001043

51. Michael CL, Mittelstaedt H, Chen Y, Desai AV, Kuperman GJ. Applying User-Centered Design in the Electronic Health Record (EHR) to Facilitate Patient-Centered Care in Oncology. AMIA Annu Symp Proc AMIA Symp. 2020;2020:833–9. PubMed PMID: 33936458; PubMed Central PMCID: PMC8075506.

52. Elwyn G, Nelson E, Hager A, Price A. Coproduction: when users define quality. BMJ Qual Saf. 2020 Sep;29(9):9. doi:10.1136/bmjqs-2019-009830

53. Vargas C, Whelan J, Brimblecombe J, Allender S. Co-creation, co-design, co-production for public health - a perspective on definition and distinctions. Public Health Res Pract. 2022;32(2):2. doi:10.17061/phrp3222211

