## Supplementary material for "Assessing EHR potential for adaptive learning in multimorbidity care in Sub-Saharan Africa: a mixed-methods study of Zimbabw’s Impilo system": S1 SRQR Checklist

### **Supporting Information 1 Checklist. Standards for Reporting Qualitative Research (SRQR) Checklist**

| **SRQR Domain** | **SRQR Item** | **Manuscript Location** | **Comments** |
| --- | --- | --- | --- |
| Title and Abstract | Title | Title Page | Identifies topic, setting, and mixed-methods design. |
| Title and Abstract | Abstract | Abstract | Summarises background, methods, findings, and conclusions. |
| Introduction | Problem Formulation | Introduction | Describes the challenge of multimorbidity care and learning-oriented EHRs in Sub-Saharan Africa. |
| Introduction | Purpose or Research Question | Introduction | States study objectives and research questions. |
| Methods | Qualitative Approach and Paradigm | Study Design | Multi-method qualitative design informed by socio-technical and Learning Health System perspectives. |
| Methods | Researcher Characteristics and Reflexivity | Study Design / Limitations | Multidisciplinary team comprising health systems researchers, clinicians, and digital health practitioners. |
| Methods | Context | Study Setting | Describes Zimbabwean health system and Impilo EHR context. |
| Methods | Sampling Strategy | Participants and Data Sources | Purposive selection of documents, facilities, patient journeys, and interview participants. |
| Methods | Ethical Issues | Ethics Statement | Ethics approvals, consent procedures, and confidentiality measures reported. |
| Methods | Data Collection Methods | Data Collection | Documentary review, ethnographic observation, patient journey mapping, and semi-structured interviews. |
| Methods | Data Collection Instruments and Technologies | Data Collection | Interview guides, observation templates, patient journey tools, and document review matrix described. |
| Methods | Units of Study | Data Sources | Documents, patient journeys, observations, and key informants. |
| Methods | Data Processing | Data Analysis | Procedures for organising, coding, and managing qualitative data described. |
| Methods | Data Analysis | Data Analysis | Matrix-based framework analysis and thematic analysis undertaken across data sources. |
| Methods | Techniques to Enhance Trustworthiness | Data Analysis | Triangulation, iterative analysis, team discussions, and comparison across data sources. |
| Results | Synthesis and Interpretation | Results | Findings organised around socio-technical domains and learning-oriented information capability. |
| Results | Links to Empirical Data | Results | Supported by documentary evidence, observations, patient journeys, and participant quotations. |
| Discussion | Integration with Prior Work | Discussion | Findings interpreted in relation to Learning Health Systems, multimorbidity, and EHR literature. |
| Discussion | Implications and Transferability | Discussion and Conclusion | Implications for EHR design, integrated care, and health system learning discussed. |
| Discussion | Limitations | Strengths and Limitations | Study limitations and strengths explicitly reported. |
| Other | Conflicts of Interest | Competing Interests | Declared in manuscript. |
| Other | Funding | Funding Statement | Funding sources and role of funders reported. |

**Researcher characteristics and reflexivity**

The study was conducted by a multidisciplinary team comprising clinicians, epidemiologists, health systems researchers, digital health practitioners, implementation scientists, and policy actors with extensive experience in HIV, multimorbidity, electronic health records, and health system strengthening in Zimbabwe. Several members of the research team had longstanding professional involvement in the development, implementation, governance, or evaluation of national digital health initiatives, including the Impilo electronic health record system and related chronic disease programmes. Consequently, many interview participants were professional colleagues, collaborators, or stakeholders with whom members of the research team had previously interacted through routine health system activities, technical working groups, programme implementation, research collaborations, or policy processes.

These existing relationships facilitated access to participants, enabled deeper contextual understanding of the health system, and supported interpretation of complex organisational and technical processes that may not have been readily apparent to external researchers. At the same time, the research team recognised that prior professional relationships, disciplinary training, and familiarity with the digital health ecosystem could shape data collection, interpretation, and the framing of conclusions. Particular attention was therefore paid to the possibility of shared assumptions regarding the strengths, limitations, and future potential of the Impilo system.

The study was undertaken within a broader programme of co-production aimed at improving the capacity of electronic health records to support multimorbidity care. Consistent with Learning Health System and realist-informed approaches, researchers did not position themselves as detached observers but as participants within a wider process of knowledge generation and system improvement. The study therefore acknowledged that findings emerged through interactions between researchers, participants, technologies, organisational structures, and the broader health system context.

To enhance reflexivity and trustworthiness, interpretations were developed iteratively through multidisciplinary discussions involving researchers with different professional backgrounds and levels of involvement in digital health implementation. Emerging findings were compared across documentary review, ethnographic observation, patient journey mapping, and interview data. Analytic memos were maintained throughout data collection and analysis to document interpretations, challenge assumptions, and support transparency in the development of findings.
