## Supplementary material for "Assessing EHR potential for adaptive learning in multimorbidity care in Sub-Saharan Africa: a mixed-methods study of Zimbabw’s Impilo system": S2 Documentary review

S4 Table: Documents included in the documentary analysis

| **Document** | **Year** | **Document category** | **Scope / analytic role** |
| --- | --- | --- | --- |
| Zimbabwe National Health Strategy | 2021–2025 | **Policy** | Sets overall health system priorities and frames expectations for service delivery within which digital health operates |
| Zimbabwe National Digital Health Strategy | 2021–2025 | **Policy** | Articulates national vision, principles, and priorities for digital health and health information systems |
| MoHCC Standard Process for Updating the Impilo EHR | 2023 | **Governance** | Defines institutional roles, approval pathways, and change-management processes governing evolution of the national EHR |
| multimorbidity in Zimbabwe: Evidence and Priorities Dialogue – Workshop Report | 2023 | **Governance, Policy, Programme** | Documents national deliberation and priority-setting on multimorbidity, synthesising evidence to inform system-level decision-making |
| Operational and Service Delivery Manual for HIV Care and Treatment (Zimbabwe) | 2022 | **Programme** | Operationalises national HIV policy, including differentiated service delivery and limited integration of selected chronic conditions |
| HIV Care and ART Clinical Record (MoHCC, Zimbabwe) | 2019 | **Programme** | Specifies the standardised HIV clinical record shaping routine data capture, longitudinal follow-up |
| Sanofi–OPHID HIV/NCD Integration Project Progress Report | 2023 | **Programme** | Provides empirical programme-level evidence on HIV–NCD screening, referral pathways, and routine data use |
| Electronic Health Record Systems in Limited Resource Settings: A Comprehensive Evaluation of the Impilo Platform | 2022 | **Empirical Research** | Provides early evaluative evidence on implementation, use, and challenges of the Impilo EHR in routine settings |
| Impilo EHR Evaluation Report | 2025 | **Evaluation** | Reports recent findings on Impilo functionality, governance, sustainability, and system integration |
| WHO Package of Essential Noncommunicable Disease Interventions (PEN) | 2020 | **Clinical guidance** | Provides normative primary-care protocols for NCD management, informing expectations for chronic disease documentation and follow-up |
| multimorbidity: A Core Priority for Learning Health Systems Amidst ‘Vertical’ Disease Programme Cuts | 2025 | **Empirical research** | Provides empirically grounded analysis of multimorbidity and learning health system constraints in Zimbabwe, situating documentary findings within observed practice |
| Multimorbidity and health system priorities in Zimbabwe | 2024 | **Empirical research** | Provides empirically grounded analysis of multimorbidity and learning health system priorities. |
