## Supplementary material for "Assessing EHR potential for adaptive learning in multimorbidity care in Sub-Saharan Africa: a mixed-methods study of Zimbabw’s Impilo system": S3 Participant summary

**Participant demographics and activity breakdown**

| Participant category | In-depth interviews | Participant observation | Patient journeys | Total |
| --- | --- | --- | --- | --- |
| People living with multimorbidity |  | 11 | 11 | **11** |
| People living with HIV only |  | 6 | 6 | **6** |
| People living with hypertension only |  | 6 | 6 | **6** |
| Health professionals (Clinics 1 and 2) | 7 | 4 |  | **11** |
| Decision makers within national MoHCC | 5 | - |  | **4** |
| Health informatics experts | 2 | - | - | **2** |
| Technical partners/NGOs | 3 | - | - | **3** |
| City Health Department (Bulawayo and Chitungwiza) | 2 | 1 | - | **3** |
| Grand total | 19 | 28 | 23 | **46** |

*Participants enrolled for patient journeys were the same ones that were observed during care.
