## Supplementary material for "Assessing EHR potential for adaptive learning in multimorbidity care in Sub-Saharan Africa: a mixed-methods study of Zimbabw’s Impilo system": S4 Patient journey

S3 Table: Characteristics of observed patient journeys, sorted by conditions present

| **Case ID** | **Sex** | **Age (years)** | **Number of LTCs** | **Observation duration^#^** |
| --- | --- | --- | --- | --- |
| PJM-01 | Female | 65+ | 1 | ~24 min |
| PJM-02 | Female | 65+ | 1 | ~38 min |
| PJM-03 | Female | 45-55 | 1 | ~1 hr 47 min |
| PJM-04 | Male | 45-55 | 1 | 60 min |
| PJM-05 | Female | 65+ | 1 | ~3 hrs |
| PJM-06 | Male | 55-65 | 1 | ~4 hrs |
| PJM-07 | Female | 35-45 | 1 | ~17 min |
| PJM-08 | Female | Unknown | 1 | 27 min |
| PJM-09 | Female | 45-55 | 2 | ~5 hrs |
| PJM-10 | Adult | 35-45 | 2 | ~4–5 hrs |
| PJM-11 | Adult | 45-55 | 2 | ~4 hrs |
| PJM-12 | Female | 55-65 | 2 | ~5 hrs |
| PJM-13 | Adult | 65+ | 2 | ~4 hrs |
| PJM-14 | Female | 65+ | 2 | ~2 hrs |
| PJM-15 | Female | 65+ | 2 | ~43 min |
| PJM-16 | Male | 35-45 | 2 | ~4–5 hrs |
| PJM-17 | Male | 45-55 | 2 | 6h 10m |
| PJM-18 | Male | 55-65 | 2 | ~7 hrs |
| PJM-19 | Female | 35-45 | 2 | ~2 hr 45 min |
| PJM-20 | Adult | 35-45 | 3 | ~5 hrs |
| PJM-21 | Female | 65+ | 3 | ~3 hrs |
